# The effectiveness of acupuncture point stimulation for the prevention of post-operative sore throat: a meta-analysis

**DOI:** 10.1101/2020.07.11.20148544

**Authors:** Pin-Yu Jau, Shang-Chih Chang

## Abstract

**Background:** Enhanced recovery pathways can be further improved for postoperative sore throat (POST) which usually occurs after surgery with general anesthesia. Medications have shown some effectiveness in treating and preventing POST, but acupuncture or related techniques with better safety and less cost likely can be used as an alternative or adjuvant therapy to treat perioperative symptoms by stimulating acupuncture point (acupoint). Therefore, we aim to conduct a meta-analysis to assess whether acupoint stimulation help patients prevent or treat POST in adults undergoing tracheal intubation for general anesthesia.

**Methods:** Publication in PubMed, the Cochrane Central Register, ScienceDirect, and ClinicalTrial.gov were surveyed from Jan. 2000 through Jan. 2020. Studies that compared intervention between point stimulation and none or sham point stimulation, were included. Primary outcomes were the incidence and severity of POST at 24h. Secondary outcomes were the incidence of postoperative nausea and vomiting, choking cough, and sputum.

**Results:** Three randomized control trials and one comparative study involving 1358 participants were included. Compared with control, acupoint stimulation was associated with a reduced incidence (risk ratio, 0.3; 95% confidence interval (CI), 0.2–0.45; *p* < 0.001) and severity (standardized mean difference, −2.21; 95% CI, −2.67 to −1.76; *p* < 0.001) of POST. Secondary outcomes are also in favor of acupoint stimulation. There were no significant adverse events related to acupoint stimulation. Subgroup, the sensitivity, and the trial sequence analyses confirmed that the finding for POST was adequate.

**Conclusions:** Acupoint stimulation with various methods may reduce the occurrence of POST. It could be considered as one of nonpharmacological ways to prevent POST in enhanced recovery pathways. Further rigorous studies are needed to determine the effectiveness of acupoint stimulation.

**Question:** Can acupoint stimulation prevent postoperative sore throat after tracheal intubation?

**Findings:** Acupoint stimulation by acupuncture or related techniques more significantly reduces the incidence and the severity of postoperative sore throat than non- /sham-treatment at 24 hours.

**Meaning:** Acupoint stimulation by acupuncture or related techniques could be an effective, nonpharmacological approach to prevent postoperative sore throat in enhanced recovery after tracheal intubation.

## INTRODUCTION

Postoperative sore throat (POST) is a common symptom caused by tracheal intubation undergoing general anesthesia^1^. The incidence of POST reported at university hospitals in Asia region varies from 35.7% to 57.5%, and the studies show higher possibility of occurrence due to some factors, such as female, older age, higher intracuff pressure, type of airway devices, duration of anesthesia, and site of surgery^2–4^. Although this postoperative symptom is often considered as a minor complication, it remains one of the most patients’ complaints after general anesthesia, second only to postoperative nausea and vomiting (PONV)^5^. In the past few years, there have been some ways to improve PONV in Enhanced Recovery after Surgery (ERAS)^6^. POST also has a chance to persist for several days and may significantly distress to the patient during postoperative care^7–8^; we believe that there are rooms for improving ERAS pathways for POST^9^.

The etiologies of throat complication are believed to be caused by tracheal intubation that results in mucosal trauma and inflammation^10–11^. Several literatures suggest some medications, such as dexamethasone^12^, benzydamine^13^, corticosteroids^14^, lidocaine^15^, ketamine^16^, and magnesium^17^, could be the effective strategies to prevent or treat POST.

In contrast to the modern treatments, acupuncture or related techniques are also used as an alternative or adjuvant therapy in perioperative time, for example, the management of postoperative pain^18^. Acupuncture is the medical practice to stimulate acupuncture points (acupoints) on the body with thin needle and improve health on the theory of traditional Chinese medicine^19^. Its related techniques, including moxibustion, acupressure, acupoint application, electrical/laser/magnetic/ultrasonic acupuncture, transcutaneous electrical nerve/acupoint stimulation (TENS/TEAS), and bloodletting, are the same in principles but different in methods^20–22^. However, the effectiveness of acupoint stimulation of various methods in order to prevent or treat POST remains inconclusive. Therefore, in this paper, the presented meta-analysis aims to examine whether acupoint stimulation help patients prevent or treat POST undergoing tracheal intubation for general anesthesia.

## METHODS

The means and the reports of this systematic review followed the Cochrane Collaboration methodology^23^ and PRISMA statement^24^. The protocol is registered at PROSPERO (CRD42020177480)

### Eligibility criteria

Regarding the types of studies, we enrolled randomized controlled trials or comparative experimental trials, and excluded follow-up studies, case series, and case reports. The target participants should have the following inclusion criteria: (1) adult patients had the elective surgery with general anesthesia; (2) all patients received intubation by various types of airway device. All retrieved studies were required to comprise at least two arms, one of which had an intervention with acupoints stimulation and the other of which had intervention with non- /sham-acupoints stimulation. Studies were also excluded if they had no evaluation of sore throat at 24h after extubation or surgery.

### Search strategy

We searched four electronic databases, including PubMed, Cochrane Central Register of Controlled Trials (CENTRAL), ScienceDirect, and ClinicalTrial.gov, in the period from January 2000 to January 2020. Original articles not written in English or Chinese and not available in full-text were excluded.

The following search terms were used individually or combined: “postoperative sore throat” ; “intubation complication” ;” anesthesia complication” ;” acupressure” ;” transcutaneous electrical nerve stimulation” ;” acupuncture” ;” acupuncture point” ;” moxibution” ;” bloodletting”. The MeSH terms were used during the search if the database system was available. Details of the search strategy was shown in Supplementary Table 1.

## Study selection

Two authors selected the included studies according to the eligibility criteria independently. Disagreement was resolved by discussion.

## Data extraction

The same authors examined the included studies and extracted data with a predetermined form. It recorded the first author, year, study design, sample size, sex, age, American Society of Anesthesiologists (ASA) physical status classification, surgery type, intervention, and outcome measurements. Detailed information about the intervention and the anesthesia was also collected.

## Risk of bias assessment

The risk of bias was assessed by the same authors independently by using the Cochrane risk of bias assessment tool^23^. To randomized trials, the RoB 2.0 tool was used to assess five domains for risk-of-bias that lead to an overall bias^25^; to non-randomized trials, the ROBINS-I tool was used to assess seven domains that may be ascertained to produce an overall bias^26^. The risk of bias was visualized by robvis^27^. Disagreement was resolved by discussion.

## Data synthesis and analysis

The primary outcome was the incidence or the severity of sore throat at 24h after surgery/extubation and adverse events in the study groups. If postoperative sore throat used traditional four-level classification system to rate the severity of the condition (i.e. none, mild, moderate, and severe), the incidence of sore throat was calculated from the sum of mild, moderate, and severe cases. The secondary outcomes included the incidence or the severity of related complications after surgery/extubation. Continuous and dichotomous outcomes were presented as risk ratios (RRs), standardized mean differences (SMDs) and 95% CIs. When trials contained zero events in either arm, continuity correction was calculated with the addition of 0.5 to each cell of 2×2 tables from the trial^28^. When continuous outcomes were presented as medians with interquartile range, they were converted to the means and standard deviation via the method proposed by Shi et al.^29^. If the data could not be applied to the meta-analysis, we summarized them in the text.

We chose DerSimonian and Laird random-effects model to analyze collected data. Heterogeneity among studies was assessed using the Higgins *I ^2^* test; *I ^2^* >50% means substantial heterogeneity. This random-effects model was often considered with inappropriate type I error and we thus conducted sensitivity analysis using Hartung-Knapp-Sidik-Jonkman method because it can work better in a limited number of included studies^30^. Subgroup analyses were also conducted to assess whether the treatment effects were changed because of study design or airway device. Trial sequential analysis was tried to perform due to examine required information size and the reliability of outcome from our meta-analysis. According to the previous review for postoperative complication and acupoint stimulation^31^, we designed a relative risk reduction of 30% for POST that was considered clinically meaningful in a 5% risk of a type I error and a power of 80%. Non-randomized studies were excluded from trial sequential analysis. Statistical significance was defined as *p* < 0.05, except for the determination of publication bias, that employed *p* < 0.1. If more than 10 studies were included in the meta-analysis, publication bias was investigated by funnel plots. Statistical analysis was conducted by Comprehensive Meta-Analysis (CMA), version 3 (Biostat, Englewood, NJ, USA) and the trial sequential analyses were performed using TSA software, version 0.9 beta (Copenhagen Trial Unit, Copenhagen, Denmark).

## RESULTS

### Overview of included studies

Our initial search yielded 130 titles and abstracts (Figure 1). After applying the inclusion and exclusion criteria, 3 randomized controlled trials and 1 quasi-experimental study involving 1358 participants were included in the analysis^32–35^. The mean age for the study participants ranged from 35.69 to 48.6 years, and the proportion of women ranged from 35.7% to 100% (Table 1). Four trials included patients with an ASA status of I–II. Three trials provided obvious type of surgery, which included total abdominal hysterectomy^32^, cholecystectomy^33^, gastrointestinal^33^, herniorrhaphy^33^, mastectomy^33^, and thyroid surgery^33–34^. One trial offered the note that all patients didn’t receive head or neck surgery^35^.

**Figure 1.**
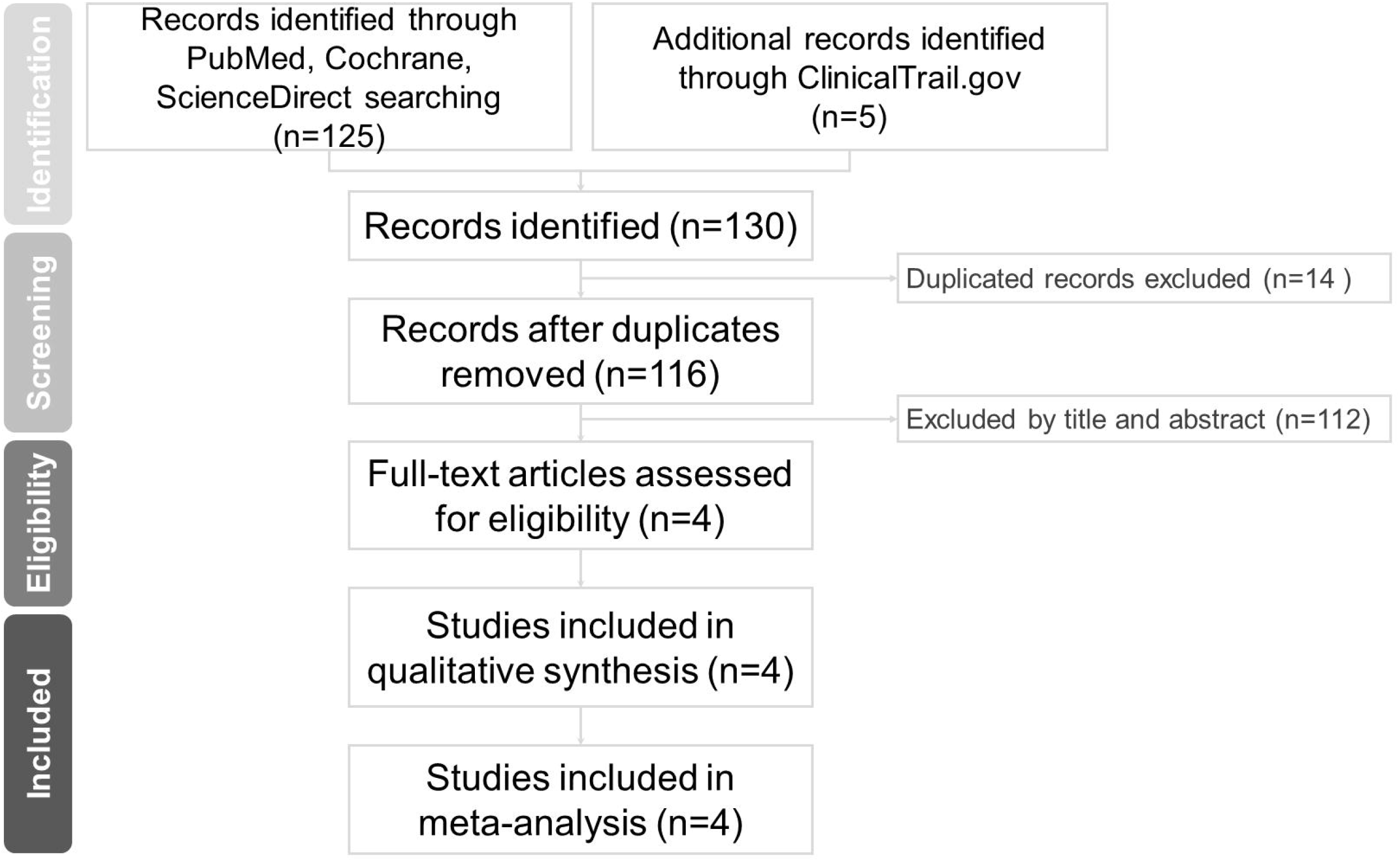
PRISMA flow diagram for searching and identification of included studies.

**Table 1.**
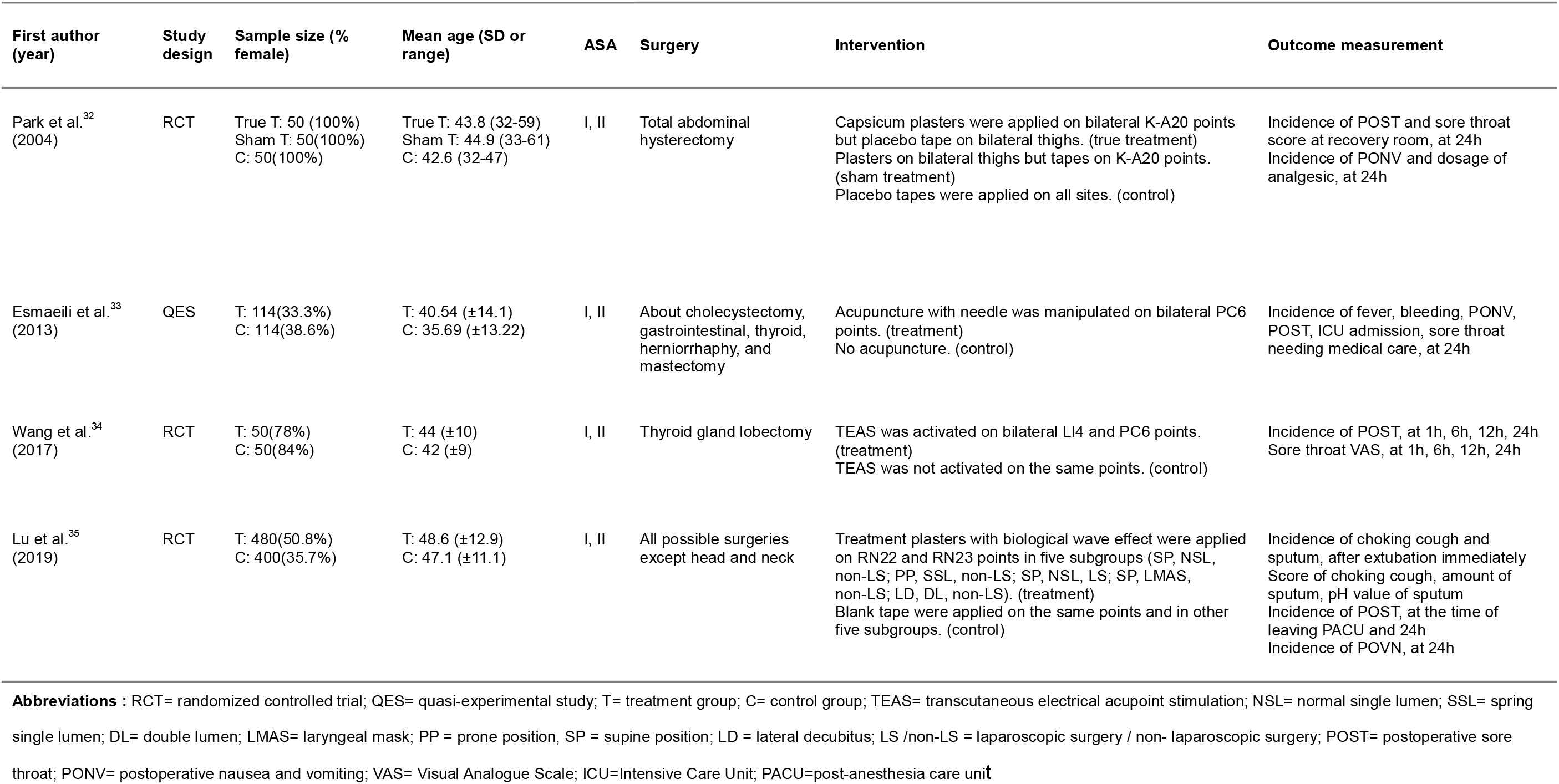
Characteristics of included studies.

One trial used capsicum plaster for acupoint application and compared true-point treatment with sham-point treatment and control^32^. Another trial used plaster with biological wave effect for acupoint application and compared true treatment with control in subgroups divided by the site of acupoint, the position of operation, the type of airway device, and the operation method^35^. The research team firstly tried to compare a 5-acupoints and 2-acupoints treatment subgroups with the control subgroup in mode of supine position, normal single lumen, non-laparoscopic surgery, and confirmed the same treatment effect between two acupoint approaches. Other treatment and control subgroups just used 2-acupoints application in other modes. We pooled data from 2-acupoints treatment subgroups in various mode to analyze because it seemed a major acupoint approach in this trial. The other trial compared the active TENS with the inactive, sham TENS^34^. The quasi-experimental study compared the case of body acupuncture with the case of no acupuncture^33^. The details of the anesthesia and the intervention were summarized in Supplementary Table 2.

One trial used a five-point scale to assess the score of sore throat^32^, and another trial used a visual analog scale (VAS) to assess the score of sore throat^34^. Besides the incidence of postoperative sore throat at 24h and the score of sore throat, there were the incidence of postoperative nausea and vomiting at 24h, the incidence of choking cough and sputum after extubation immediately in outcome which could be available to analyze the effectiveness of acupoint stimulation.

### Risk of bias

The quality of randomized control trials generally needed to have some concerns according to the RoB 2.0 (Figure 2a). Two included studies had unclear random sequence generation and allocation concealment^34–35^; one of them didn’t report enough information how to blind when participants received intervention before anesthesia induction^34^. The quality of quasi-experimental study also needed some concerns according to the ROBINS-I (Figure 2b). Although all participants in study received tube and had intubation-related trauma, other possible undetermined causes of sore throat could generate confounding factors and could lead to a vague outcome of selection or measurement^33^.

**Figure 2.**
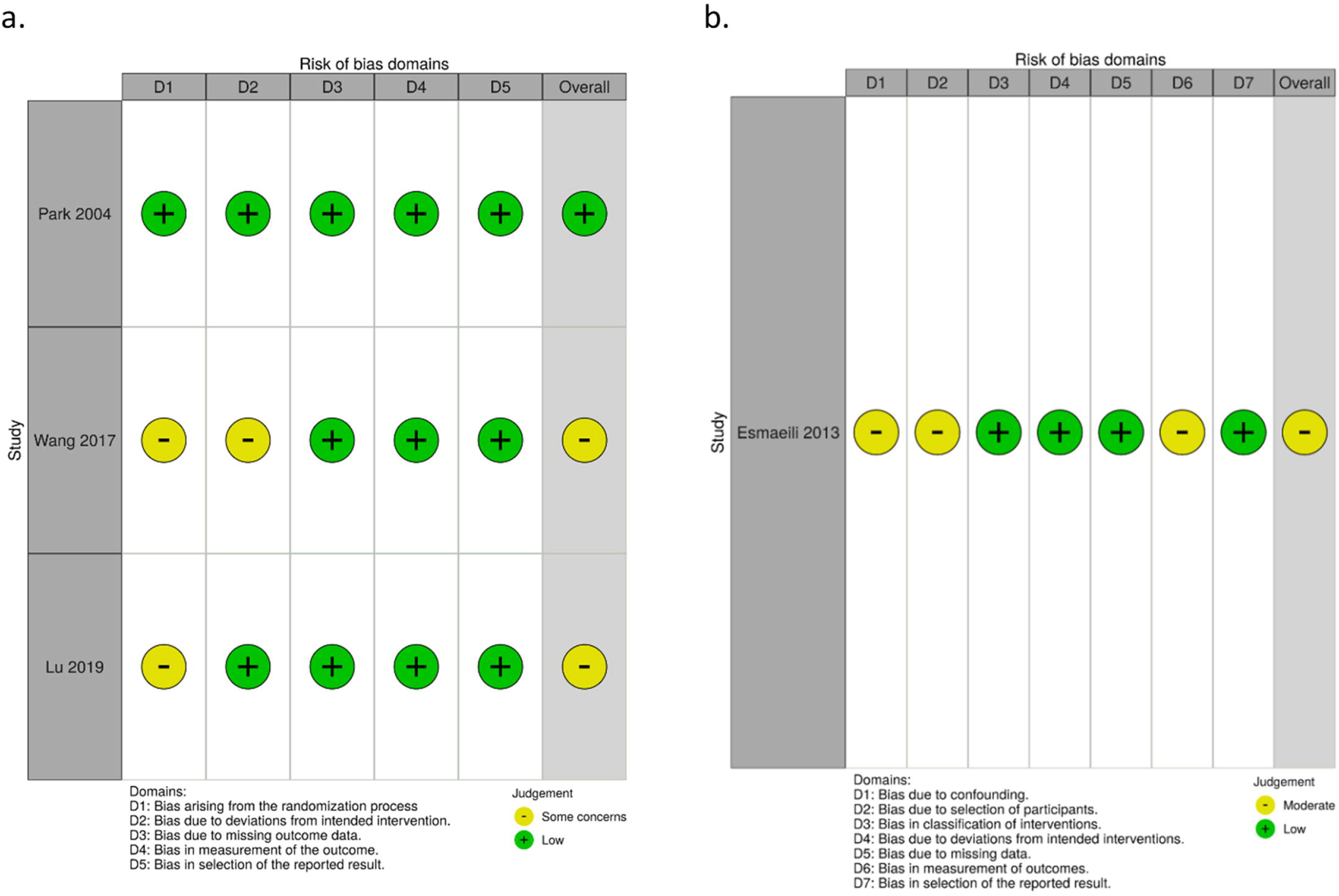
Risk of bias summery for included studies. (a) 3 RCT via RoB 2.0; (b)1 QES via ROBINS-I. RCT: randomized controlled trial; QES: quasi-experimental study.

### Primary outcomes

#### Incidence of postoperative sore throat

Four studies that included 1228 participants provided data on the incidence of sore throat at 24h after surgery or extubation^32–35^. The resulted indicated that acupoint stimulation was associated with a reduced incidence of postoperative sore throat (RR, 0.3; 95% CI, 0.2– 0.45; *p* < 0.001; df=7; *I*^2^=0; Figure 3a). The number needed to prevent postoperative sore throat was 6 (95% CI, 6–7).

**Figure 3.**
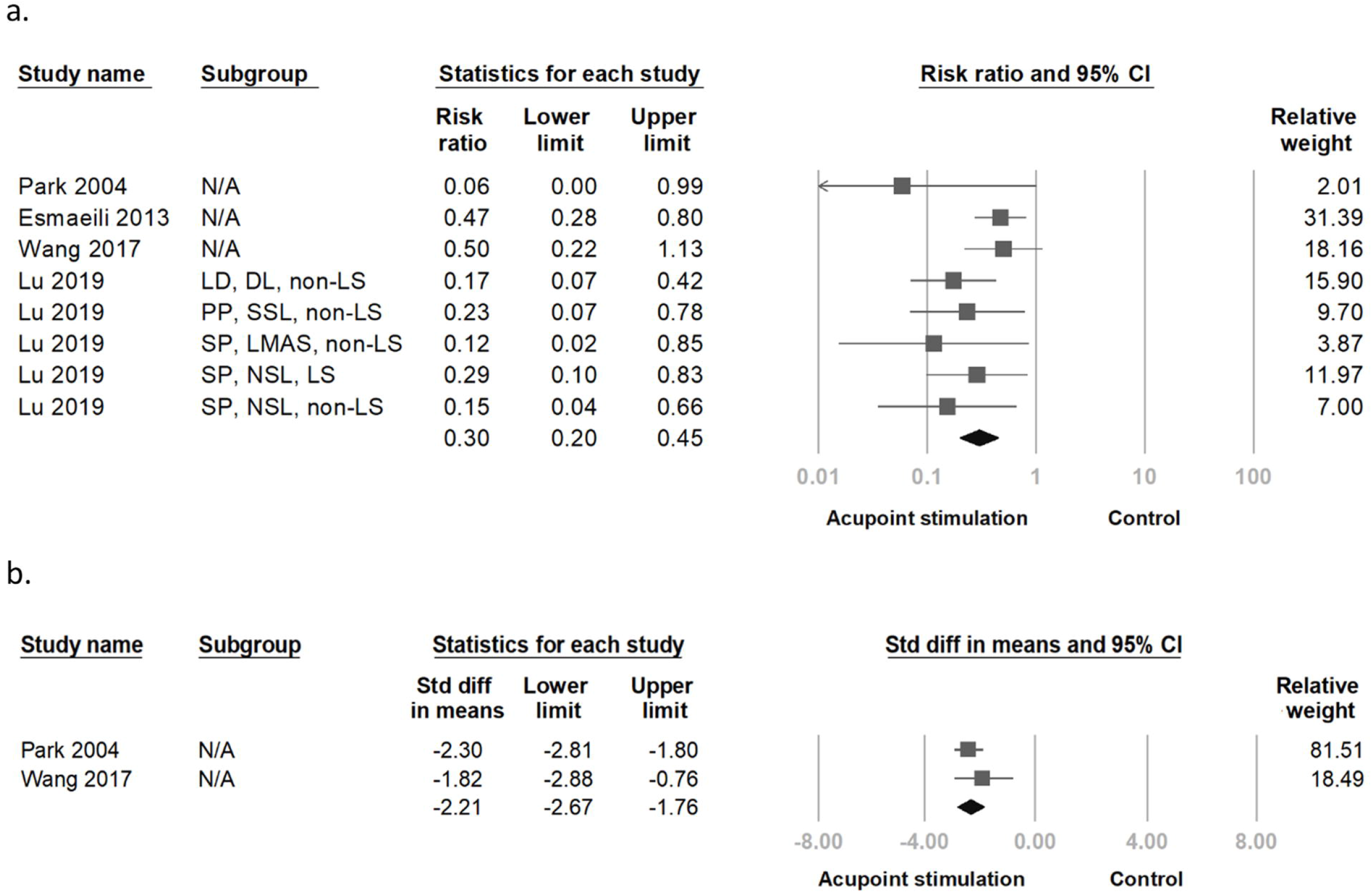
The forest plot presented (a) the incidence and (b) the severity of postoperative sore throat at 24h in patients who received the intervention between point stimulation and none / sham point stimulation. NSL: normal single lumen; SSL: spring single lumen; DL: double lumen; LMAS: laryngeal mask; PP: prone position, SP: supine position; LD: lateral decubitus; LS /non-LS: laparoscopic surgery / non-laparoscopic surgery

#### Severity of postoperative sore throat

Two studies involving 121 participants offered the severity of sore throat at 24h after surgery or extubation^32,34^. One study assessed the postoperative sore throat at 24h with VAS mean score and standard deviation (SD) that had significantly difference between the true treatment and the sham treatment via independent t-test^34^; the other reported the sore throat scores at 24h with median and range that were significantly lower in the true-point treatment group than sham-point treatment and control via Kruskal-Wallis test^32^. However, by converting median (range) to mean (SD) in math and pooled analyzing, we found that the acupoint stimulation was associated with decreased severity of postoperative sore throat (SMD, -2.21; 95% CI, −2.67 to −1.76; *p* < 0.001; *I*^2^=0; Figure 3b).

#### Adverse events

One study reported that two patients in the true-point treatment and four patients in the sham-point treatment had the plaster side-effect about mild burning sensation with erythema^32^. The other didn’t reported any adverse events developed in their study groups^33–35.^

### Secondary outcomes

#### Incidence of postoperative nausea and vomiting

Three studies that included 1128 participants provided data on the incidence of postoperative nausea and vomiting at 24h after surgery or extubation^32,33,35^. The trial offered the incidence of nausea and vomiting individually, but we chose the incidence of nausea to analyze because less commonly vomiting occurred without nausea in adult and the proportion of postoperative nausea and vomiting could approximate it^32^. The result showed that the acupoint stimulation was associated with a reduced incidence of postoperative nausea and vomiting (RR, 0.49; 95% CI, 0.3–0.79; *p* = 0.003; df=6; *I*^2^=0; Figure 4a).

**Figure 4.**
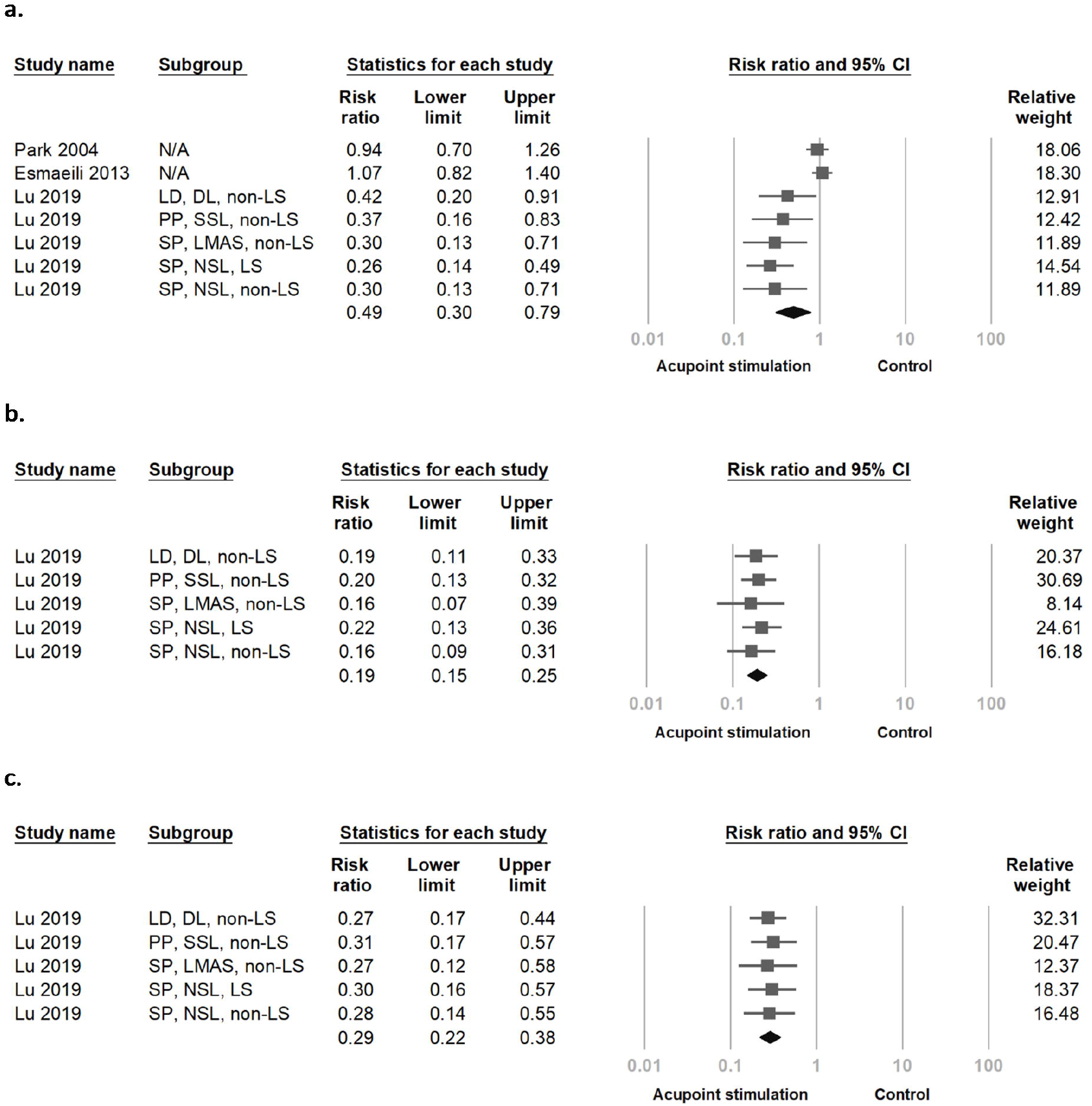
The forest plot presented the incidence of (a) postoperative nausea and vomiting at 24h and the incidence of (b) choking cough, (c) sputum immediately after exbutaion in patients who received the intervention between point stimulation and none / sham point stimulation. NSL: normal single lumen; SSL: spring single lumen; DL: double lumen; LMAS: laryngeal mask; PP: prone position, SP: supine position; LD: lateral decubitus; LS /non-LS: laparoscopic surgery / non-laparoscopic surgery

#### Incidence of choking cough and sputum

The studies with 800 participants provided data on the incidence of choking cough and sputum immediately after extubation^35^. The acupoint stimulation was associated with a reduced incidence of choking cough (RR, 0.19; 95% CI, 0.15–0.25; *p* < 0.001; df=4 ; *I*^2^=0; Figure 4b) and a reduced incidence of sputum (RR, 0.29; 95% CI, 0.22–0.38; *p* < 0.001; df=4 ; *I*^2^=0; Figure 4c).

### Subgroup, sensitivity, and trial sequential analyses

In subgroup analysis, for study design, we divided four studies into RCT and QES group. It was no significant difference between two groups (Q-value, 3.099; *p* =0.078; df=1), but pooled analysis with only RCT group could show more reduced incidence of POST (RR, 0.25; 95% CI, 0.16–0.39; *p* < 0.001). For airway device, we inferred that three studies used single lumen tube from the description of anesthesia process, tube size and surgery type in the articles^32–34^, and divided four studies into single lumen, double lumen, and laryngeal mask group. There was no significant difference among three groups (Q-value, 3.35; *p*=0.187; df=2) and all tended to favor acupoint stimulation for reducing the incidence of POST.

We conducted sensitivity analysis on the incidence of POST, PONV, choking cough and sputum via Hartung-Knapp-Sidik-Jonkman method. The aggregate effect in each outcome was consistent with the primary finding and all were still statistically significant despite becoming larger range of 95% CI (Supplementary Table 3). When excluding QES, trial sequential analysis with RCTs suggested that the cumulative z-curve crossed both the conventional and trial sequential monitoring boundaries for benefit before reaching the required information size (1602 patients), thereby recommending a positive effect of acupoint stimulation on the prevention of postoperative sore throat (Supplementary Figure 1).

## DISCUSSION

According to limited data derived from the included trials, our study suggests that acupoint stimulation in adults undergoing tracheal intubation for surgery under general anesthesia is associated with a decline of POST at 24h and possibly results in less severity in comparison with non- / sham-acupoint stimulation. It seems a good prophylactic effect that the number needed to prevent postoperative sore throat is 6. A decline of PONV in our study is consistent with the current evidence-based article^31^. Both of decline of choking cough and sputum in our study also provide the likely positive effect when facing complications related to the removal of airway device. Limited evidence suggests that acupoint stimulation is not related with significant negative-events. Although we are not able to collect studying samples with the best quality for meta-analysis, our findings are robust throughout subgroup, sensitivity, and trial sequential analyses, thereby confirming that acupoint stimulation prevents postoperative sore throat.

As normal, the number of patients with POST decrease within 48 h and a few patients still have symptom at 96h^8^. To improve POST, most current medications are assessed via the incidence / severity of POST at 24h^12–17^. The possible mechanisms of medications to prevent or treat POST are anti-inflammatory effect, sensory-nerves suppression, and N-methyl-D-aspartic acid (NMDA) receptor antagonists^12–17^. In our view, the possible mechanisms of acupoint stimulation to prevent or treat POST are two proposed possibilities. One is to regulate anti-inflammatory response, increase local blood circulation and reduce reginal inflammatory-related pain^36–37^. The other is to start the neurophysiological system and induce acupuncture-analgesic effect^38^.

Although the clinical application of acupoint stimulation in POST remains less, previous reviews in related fields are encouraging. Lu et al.^39^ suggests that the perioperative period under anesthesia combined with acupuncture might be benefit to patients with less analgesic consumption, lower incidence of complications and better recovery; the article also gives some indications, such as appropriate acupoints selection, electric stimulation with better analgesia, and targeted population. Yoo et al.^40^ shows that enhanced recovery in gynecological surgery with acupuncture improves gastrointestinal motility, coldness, PONV, sore throat, and urinary retention. ERAS pathways still require multimodal analgesia to reduce pain, improve analgesic-related adverse effects and accelerate postsurgical recovery^41^. Acupoint stimulation by acupuncture and related techniques seems to be an effective, nonpharmacological approach with better safety and less cost in favor of ERAS^20, 39, 42–43^. Yuan et al.^44^ also proposes the concept of “perioperative acupuncture medicine”, which is the intervention of acupuncture or related techniques with acupoint stimulation before, during and after surgery, and hopefully it could be developed within ERAS.

There are a few limitations in this review. Firstly, we enroll non- / low-quality randomized trials to analyze and they limit the strength of our findings. Secondly, there is not enough sample size to decide the effectiveness of our finding. Thirdly, only one study mentions adverse events. Finally, our search is limited to English and Chinese articles only, causing a potential language bias.

In summary, true treatment with acupoints stimulation is significantly more effective than non- /sham-treatment for the prevention of postoperative sore throat at 24h. Our finding also shows more positive result about POVN, choking cough, and sputum when stimulating acupoints. Acupoints stimulation could be considered as one of nonpharmacological ways to prevent POST in ERAS. However, it still needs further clinical trials to determine the effectiveness of acupoint stimulation and adverse events due to lack of low bias risk and high-quality studies.

## Data Availability

The data that support the findings of this study are available on request from the corresponding author.

## Glossary of Terms

RCT: randomized controlled trial
QES: quasi-experimental study
TENS: transcutaneous electrical nerve stimulation
TEAS: transcutaneous electrical acupoint stimulation
NSL: normal single lumen
SSL: spring single lumen
DL: double lumen
LMAS: laryngeal mask
PP: prone position
SP: supine position
LD: lateral decubitus
LS /non-LS: laparoscopic surgery / non-laparoscopic surgery
POST: postoperative sore throat
PONV: postoperative nausea and vomiting
ERAS: Enhanced Recovery after Surgery
VAS: Visual Analogue Scale
ICU: Intensive Care Unit
PACU: post-anesthesia care unit

---

**Supplemental Table 1**. Table that demonstrates the search strategy.

**Supplemental Table 2**. Table that presents the detail information of anesthesia and intervention.

**Supplemental Table 3**. Table that demonstrates the sensitivity analysis via Hartung-Knapp-Sidik-Jonkman method.

**Supplemental Figure 1**. Figure that demonstrates the trial sequential analysis with median control event proportion of 16%.

## Notes

**Conflicts of interest:** None

### Competing Interest Statement

The authors have declared no competing interest.

### Funding Statement

The author(s) received no specific funding for this work.

### Author Declarations

Reviews, meta-analyses, or descriptions of educational materials do not involve human subjects and do not require IRB review. <Sullivan GM. Irb 101. J Grad Med Educ. 2011;3(1):5-6. doi:10.4300/JGME-D-11-00005.1>

